# Mental health outcomes of encephalitis: an international web-based study

**DOI:** 10.1101/2023.02.03.23285344

**Authors:** Yasmin Abdat, Matt Butler, Michael Zandi, Benedict D Michael, Ester Coutinho, Timothy R Nicholson, Ava Easton, Thomas A Pollak

## Abstract

Encephalitis is associated with psychiatric symptoms in the acute and post-acute stages, and many survivors experience long-term sequelae. Despite this, the breadth and severity of mental health symptoms in survivors of encephalitis has not been systematically reported. We recruited adults who had been diagnosed with encephalitis of any aetiology to complete a web-based questionnaire assessing a wide range of mental health symptoms and disorders. In total, 445 respondents from 31 countries (55.1% UK, 23.1% USA, 2.2% low-and middle-income countries) completed the survey, with a median seven years since encephalitis diagnosis; 84.7% were diagnosed by a neurologist or infectious diseases doctor. Infectious encephalitis constituted 65.4% of cases, autoimmune 29.7%. Mean age was 50.1 years (SD 15.6); 65.8% were female. The most common self-reported psychiatric symptoms were anxiety (75.2%), sleep problems (64.4%), mood problems (62.2%), unexpected crying (35.2%), and aggression (29.9%). Rates of self-reported psychiatric diagnoses following encephalitis were high: anxiety (44.0%), depression (38.6%), panic disorder (15.7%), and post-traumatic stress disorder (PTSD, 21.3%); these rates were broadly consistent with the results of a validated self-report measure, the Psychiatric Diagnostic Screening Questionnaire (PDSQ). Severe mental illnesses such as psychosis (3.3%) and bipolar affective disorder (3.1%) were also reported. Many respondents also felt they had symptoms of disorders including anxiety (37.5%), depression (28.1%), PTSD (26.8%), or panic disorder (20.9%) which had not been diagnosed by a professional. Overall, rates of major self-reported psychiatric diagnoses and symptoms did not significantly differ between autoimmune and infectious encephalitis. In total, 37.5% of respondents had thought about suicide, and 4.4% had attempted suicide since their encephalitis diagnosis. Over half (53.5%) reported that they either had no, or substandard, access to appropriate care for their mental health. High rates of sensory hypersensitivities (>75%) suggest a previously unreported association between encephalitis and this distressing symptom cluster. This large international survey indicates that psychiatric symptoms following encephalitis are common, and that mental healthcare provision to this population may be inadequate, highlighting a need for increased provision of proactive psychiatric care for these patients.

## Introduction

Encephalitis is a devastating disorder with a likely global incidence of 4.3-7.3/100,000 per year [1]. It is defined by the presence of inflammation in the brain parenchyma associated with clinical evidence of neurological dysfunction [2]. Known aetiologies are grouped into two categories: infectious and immune-mediated [3]. For as many as 37% of patients, the cause is unknown, although improving diagnostics may be reducing this figure [4].

The clinical presentations of acute encephalitis differ and are dependent on factors such as the causative agent and the brain regions affected. Common features of encephalitis include headache, seizures, focal neurological signs, movement disorders, personality/behavioural changes, and cognitive impairment/confusion [4]. Limbic encephalitis typically features prominent cognitive and behavioural symptoms in the acute phase, consistent with the known functions of the limbic system, but focal encephalitides and panencephalitides may also present with these features.

Although direct comparisons of the frequency of psychiatric presenting features are lacking, it is generally agreed that autoimmune encephalitis is more likely than infectious encephalitis to present with isolated psychiatric features or more prominent psychiatric features in the context of subtle neurological signs. In an early series of NMDA receptor antibody encephalitis, as many as 80% of patients were initially assessed by mental health services [5]. This figure has decreased as awareness of the condition has improved [6] but misdiagnosis of all kinds of encephalitis still occurs [7].

Mortality estimates vary by geography and aetiology. In infectious encephalitis mortality can be up to 30% with treatment depending on cause, while in autoimmune encephalitis mortality ranges between 12% and 40% [1]. Many more patients survive but are left with life-changing neurological injury and dysfunction. Several studies have found that survivors of encephalitis commonly experience sequelae including physical, cognitive, emotional, behavioural, and social difficulties [8–10].

While cognitive sequelae have been explored in some detail, far fewer studies have directly addressed mental health outcomes [11, 12]. Of these, the majority have had small sample sizes or focus on a relatively narrow range of mental health symptoms [13–18]. Furthermore, much research focuses on specific subtypes of encephalitis in specific population groups (e.g. children) and therefore may be unrepresentative. For these reasons, further research is required to assess, in greater detail, the mental health outcomes experienced in those who have had encephalitis of all aetiologies.

To date, there have been no large-scale surveys from the patient’s perspective exploring the long-term mental health outcomes in adults who have been diagnosed with encephalitis. We aimed to explore the prevalence and the breadth of self-reported mental health outcomes in this population. We aimed to collect data on symptoms as well as self-reported diagnoses, as well as using a validated diagnostic screening tool for common mental health disorders. We also wished to better understand patients’ experiences of their initial diagnosis, the management of their post-encephalitis symptoms, and perceptions about the impact of their encephalitis diagnosis. To achieve this, we created a web-based questionnaire which was distributed to an international sample of encephalitis patients.

## Methods

### Design and materials

This was a cross-sectional observational study. A web-based questionnaire was created using the platform Qualtrics. Respondents were recruited through an open access link via social media platforms and mailing lists of the Encephalitis Society for 19 weeks. Questions were written in clear English, and where possible, medical terminology was avoided. Support from the Encephalitis Society in the design of the study was received. The questionnaire can be found in Supplementary Information. Respondents were individuals who self-reported a diagnosis of encephalitis, of any subtype, by a medical professional.

Respondents were asked to read the electronic participant information sheet which detailed the aims and scope of the study. Following this, respondents were asked to provide their consent for their data to be submitted anonymously. It was made clear to respondents that they could withdraw from the study at any point. A single £50 voucher prize draw was offered to respondents who agreed to provide their email address (which was kept separate from the questionnaire data).

Question themes included basic demographics, encephalitis diagnosis and characteristics, as well as experience with and management of post-encephalitis symptoms/diagnoses, and illness perceptions. Questions were in the format of tick-boxes, visual analogue scales (VAS), and free-text fields. Where respondents were asked to rate the degree to which they agree to a statement, this was on a scale from ‘0’ representing strongly disagree’ to ‘50’ representing neither agree nor disagree’ to ‘100’ representing strongly agree.

Questions on mood were adapted from the Maudsley three item VAS (M3VAS) which has been validated against the Quick Inventory of Depressive Symptomatology 16-item scale [19]. Respondents were, for example, asked to rate their current mood over the past 2 weeks on a visual scale of 1–100, where 0 represented ‘Not at all depressed’, and 100 represented ‘extremely depressed’.

Respondents were asked questions from the Brief Illness Perception Questionnaire, a validated scale used to assess the cognitive and emotional representations of illness [20]. In this study, questions were modified to change the word ‘illness’ to ‘post-encephalitis symptoms’, and respondents were asked to rank each answer on a VAS ranging from, for example, 0 = no control at all to 100 = full control.

Respondents completed the Psychiatric Diagnostic Screening Questionnaire (PDSQ), a brief, self-report scale designed to screen for the 13 of the most common mental health disorders as per the Diagnostic and Statistical Manual of Mental Disorders: DSM-IV [21]. The PDSQ responses were scored as per the PDSQ scoring instructions.

### Data analysis

Data were analysed using IBM SPSS Statistics 28. Age was calculated as year of birth to survey year; similarly, length of symptoms and time since diagnosis were calculated as year of occurrence to survey year (2022). Unless otherwise stated, data is presented as mean (± standard deviation) or median (IQR). Where respondents were given the option to select more than one answer, mutually incompatible answers were removed from analysis.

Descriptive statistics (i.e., frequencies, proportions/percentages, measures of central tendency and dispersion) were used to summarize the data. Chi-square, independent t-tests, and ANOVA tests were used where appropriate.

### Ethics

This study conforms with the World Medical Association Declaration of Helsinki. The study was approved by King’s College London Research Ethics Committee (REF: 17778).

## Results

### Demographics

In total, 445 respondents from 31 countries completed the survey (2.2% from low-and middle-income countries), *Figure 1*. All 445 respondents reported that they had been diagnosed with encephalitis by a medical professional. Of those that had been diagnosed with encephalitis, 300 (72.8%) reported that they had been diagnosed by a neurologist, 49 (11.9%) by an infectious diseases doctor, nine (2.2%) by a paediatrician, two (0.5%) by a psychiatrist, and 52 (12.6%) by another type of doctor.

**Figure 1:**
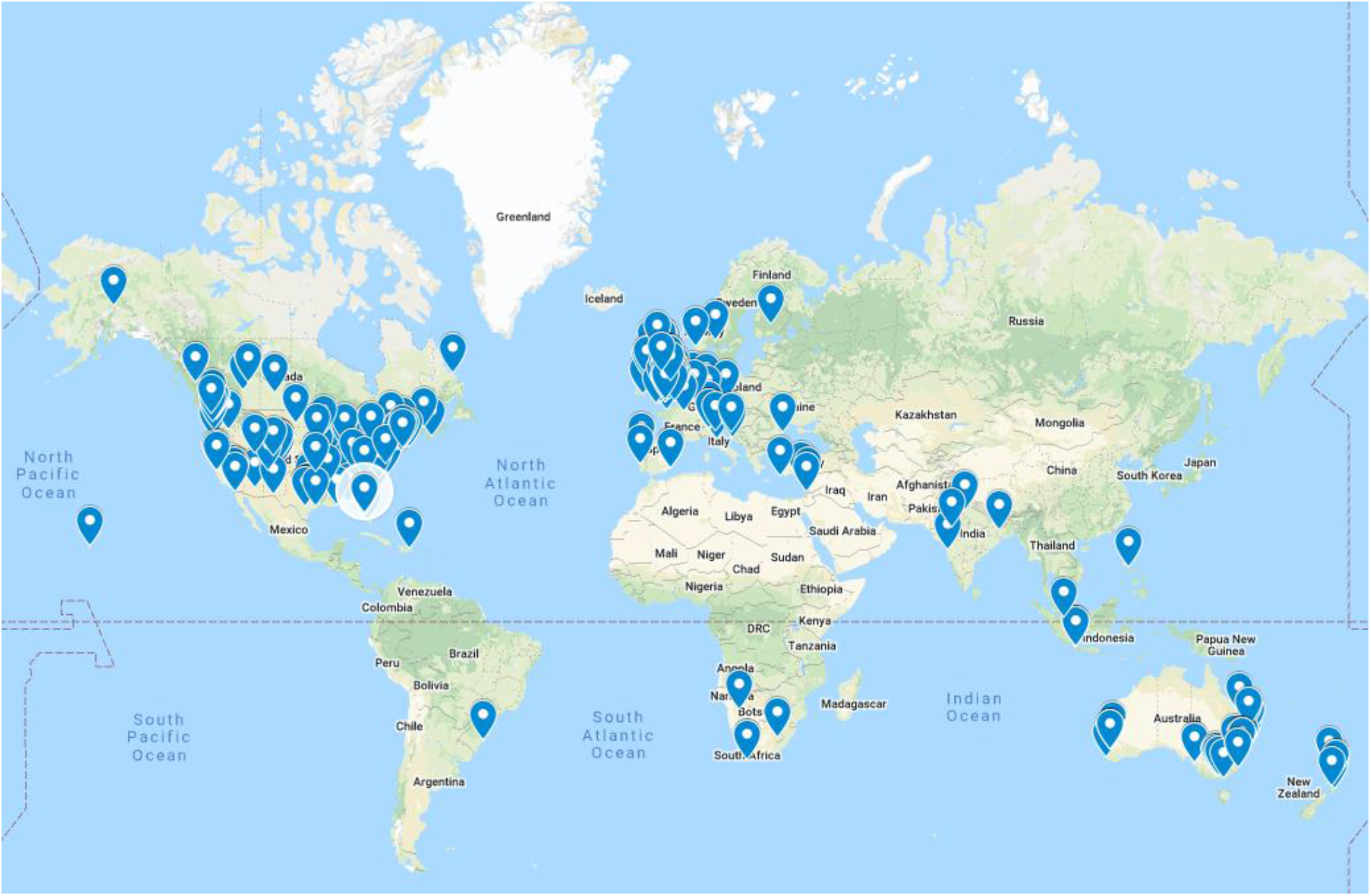
Global distribution of respondents.

The mean age of respondents was 50.1 years (SD 15.6), ranging from 18 to 85 years of age; 292 (65.8%) were female. Most respondents resided in the United Kingdom (n = 245, 55.1%) and the United States of America (n = 103, 23.1%,), but responses were received from 29 additional countries (*Figure 1)*. In total, 432 (98.2%) had completed mandatory education. Over half of respondents (n = 241, 54.3%,) were married. A range of current employment statuses were reported: 182 (40.9%) were in some form of unemployment with over a quarter unemployed (n = 122, 27.4%).

### Acute encephalitis

Average time since symptoms began was median 7.0 years (IQR 3.0-15.0 years). Infectious encephalitis constituted 286 (65.4%) cases; the most common causative infectious agent was herpes simplex virus (HSV; n = 167, 41.0% of all respondents). Autoimmune encephalitis constituted 130 (29.7%) cases with the most common type being anti-NMDAR encephalitis (n = 38, 9.3% of all respondents), *Figure 2*.

**Figure 2:**
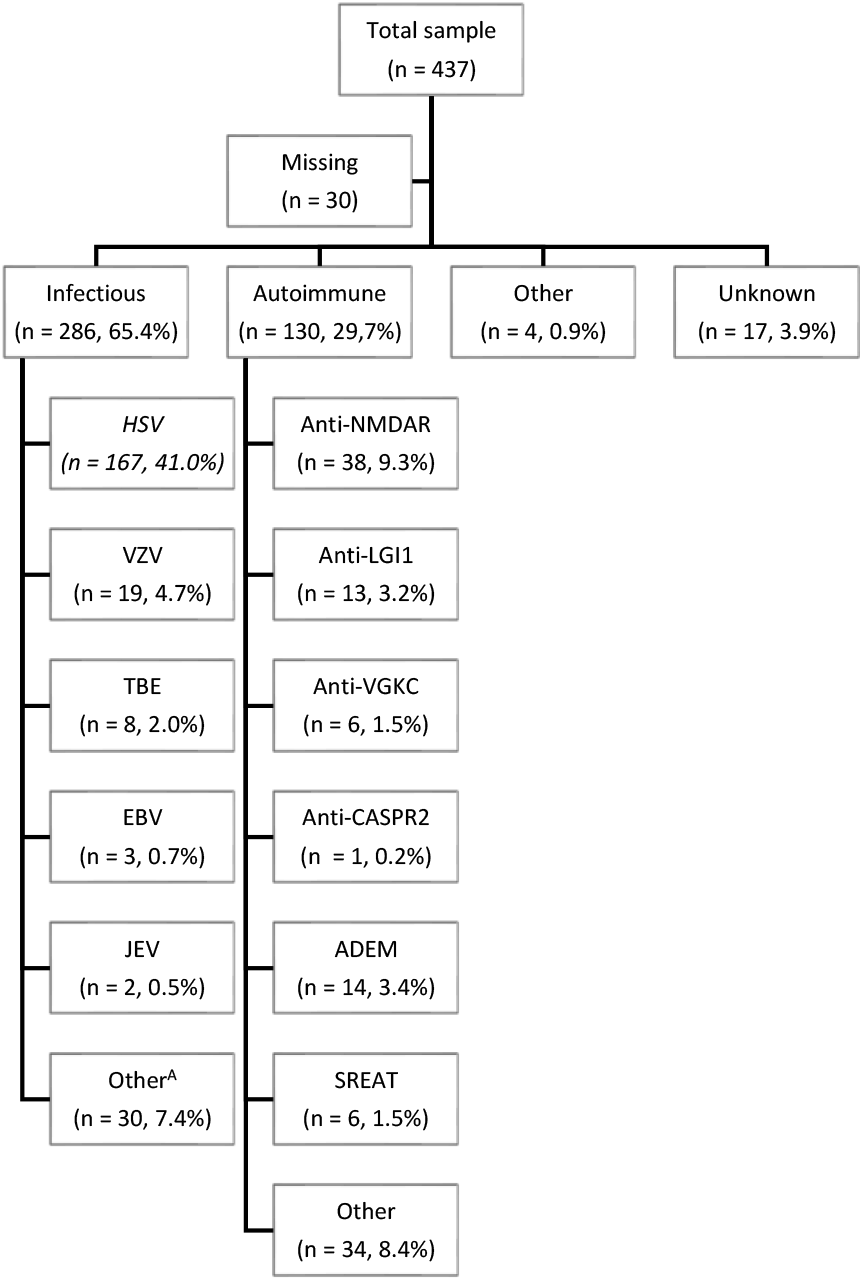
Diagnosis of encephalitis in respondents. Not all respondents (n = 437) answered questions on the aetiological agent (n = 407). A: Where known, other aetiologies included: mumps (n = 4), influenza (n = 2), COVID-19 (n = 1), equine (n = 1), measles (n = 1), mycoplasma (n = 1), scarlet fever (n = 1), West Nile virus (n = 1). B: Where known, other aetiologies included: ADEM (n = 1), Bickerstaff (n = 1), acute necrotising encephalopathy (n = 1), anti-Dipeptidyl-peptidase-like protein-6 (n = 1), anti-GAD (n = 1), anti-glial fibrillary acidic protein (n = 1), anti-ganglionic acetylcholine receptor (n = 1), anti-Hu (n = 1), CNS vasculitis (n = 1), paraneoplastic (n = 1), Rasmussen (n = 1).

Nearly half of all respondents (n = 209, 47.5%) reported having received the incorrect diagnosis prior to the correct encephalitis diagnosis. Incorrect diagnoses amongst all respondents included physical diagnoses (n = 131, 29.8%), psychiatric diagnoses (n = 37, 8.4%), both psychiatric and physical diagnoses (n = 15, 3.4%), and unknown (n = 5, 1.1%). The most commonly reported misdiagnoses (n = 200) included upper respiratory tract infection (n = 24, 12.0%), meningitis (n = 17, 8.5%), migraine (n = 13, 6.5%), mixed psychiatric (n = 12, 6.0%), mixed medical (n = 11, 6.1%) and psychiatric not otherwise specified (n = 11, 6.1%).

#### Severity of acute illness

Of valid responses (n=442), 268 (60.6%) reported being admitted to a general hospital ward, 40 (9.0%) to intensive care or high dependency care, and 53 (12.0%) reported they were not admitted to hospital. Thirty respondents (6.8%) were admitted to a psychiatric hospital. Overall, there was no effect of severity (based on admission type) on likelihood of developing a psychiatric outcome.

### Psychiatric symptoms and diagnoses

Self-reported psychiatric symptoms following respondents’ encephalitis diagnosis are presented in *Table 2*. In total, 402 (90.4%) respondents indicated that they had at least one current psychiatric symptom. Self-reported psychiatric diagnoses made by a medical professional are presented in *Table 3*. Numbers of respondents that reported they were suffering from a psychiatric diagnosis that has NOT been formally diagnosed by a healthcare professional are reported in *Table 4*

**Table 1:** Demographics of respondents from the study.^a^ Two preferred to self-describe, one preferred not to say ^b^ ‘other’ includes; Belgium (1), Brazil (1), Croatia (2), Cyprus (1), Dominican Republic (1), Finland (1), France (1), Germany (3), Greece (1), India (4), Indonesia (3), Ireland (6), Israel (1), Italy (4), Lebanon (1), Lesotho (1), Namibia (1), Netherlands (1), New Zealand (5), Norway (2), Philippines (1), Portugal (2), Romania (1), Singapore (1), South Africa (1), Spain (2), Switzerland (2).

|  | **N** | **%** |
| --- | --- | --- |
| **Gender<sup>a</sup>** | **444** | **-** |
| *Female* | 292 | 65.8% |
| *Male* | 149 | 33.6% |
| **Age** | **444** | **-** |
| *Mean (SD)* | 50.1 (SD: 15.6) | - |
| *Median* | 51.0 | - |
| *Range* | 67 | - |
| *Minimum age* | 18 | - |
| *Maximum age* | 85 | - |
| **Level of education** | **440** | **-** |
| *Did not complete mandatory education* | 8 | 1.8% |
| *Completed mandatory education* | 104 | 23.6% |
| *Completed higher education* | 109 | 24.8% |
| *Completed tertiary education/university* | 219 | 49.8% |
| **Relationship status** | **444** | **-** |
| *Single* | 107 | 24.1% |
| *In a relationship* | 27 | 6.1% |
| *Married* | 241 | 54.3% |
| *Divorced/separated* | 42 | 9.5% |
| *Other* | 27 | 6.1% |
| **Employment status** | **445** | **-** |
| *Full-time* | 97 | 21.8% |
| *Part-time* | 55 | 12.4% |
| *Self-employed* | 30 | 6.7% |
| *Unemployed* | 122 | 27.4% |
| *Retired* | 95 | 21.3% |
| *Student* | 20 | 4.5% |
| *Other* | 26 | 5.8% |
| **Country of residence** | **445** | **-** |
| *United Kingdom and Northern Ireland* | 245 | 55.1% |
| *United states of America* | 103 | 23.1% |
| *Australia* | 34 | 7.6% |
| *Canada* | 12 | 2.7% |
| *Other<sup>b</sup>* | 51 | 11.5% |

**Table 2:** breakdown of the symptoms experienced by respondents in the months or years following encephalitis diagnosis.

| Symptom | N | Yes, currently and in the past | Yes currently, but not in the past | Yes, In the past but not currently | No |
| --- | --- | --- | --- | --- | --- |
| Hallucinations | 412 | 27 (6.6%) | 11 (2.7%) | 125 (30.3%) | 249 (60.4%) |
| Paranoia | 405 | 53 (13.1%) | 12 (3.0%) | 94 (23.2%) | 246 (60.7%) |
| Mood problems | 426 | 222 (52.1%) | 43 (10.1%) | 84 (19.7%) | 77 (18.1%) |
| Aggression/anger management problems or violence | 414 | 90 (21.7%) | 34 (8.2%) | 83 (20.0%) | 207 (50.0%) |
| Anxiety | 428 | 268 (62.6%) | 54 (12.6%) | 52 (12.1%) | 54 (12.6%) |
| Inappropriate or unexpected laughter | 412 | 37 (9.0%) | 12 (2.9%) | 60 (14.6%) | 303 (73.5%) |
| Inappropriate or unexpected crying | 415 | 110 (26.5%) | 36 (8.7%) | 73 (17.6%) | 196 (47.2%) |
| Sleep problems | 427 | 223 (52.2%) | 52 (12.2%) | 60 (14.1%) | 92 (21.5%) |

**Table 3:** Formal mental health or psychiatric diagnoses in respondents

| **Diagnosis** | **N** | **Yes currently** | **Yes, in the past but after my encephalitis diagnosis** | **Yes, in the past but before my encephalitis diagnosis** | **No** |
| --- | --- | --- | --- | --- | --- |
| Depression | 409 | 101 (24.7%) | 57 (13.9%) | 34 (8.3%) | 217 (53.1%) |
| Anxiety | 402 | 127 (31.6%) | 50 (12.4%) | 35 (8.7%) | 190 (47.3%) |
| Panic disorder | 396 | 32 (8.1%) | 30 (7.6%) | 15 (3.8%) | 319 (80.6%) |
| Post-traumatic stress disorder | 396 | 43 (10.9%) | 41 (10.4%) | 14 (3.5%) | 298 (75.3%) |
| Obsessive compulsive disorder | 397 | 25 (6.3%) | 15 (3.8%) | 7 (1.8%) | 350 (88.2%) |
| Psychotic disorders | 400 | 7 (1.8%) | 6 (1.5%) | 9 (2.3%) | 378 (94.5%) |
| Bipolar disorder | 398 | 9 (2.3%) | 3 (0.8%) | 6 (1.5%) | 380 (95.4%) |
| Personality disorder | 398 | 18 (4.5%) | 13 (3.3%) | 10 (2.5%) | 357 (89.7%) |
| Impulse control disorder | 397 | 8 (2.0%) | 13 (3.3%) | 1 (0.3%) | 375 (94.5%) |
| Alcohol dependence | 398 | 7 (1.8%) | 7 (1.8%) | 5 (1.3%) | 379 (95.2%) |
| Drug dependence | 397 | 7 (1.8%) | 2 (0.5%) | 4 (1.0%) | 384 (96.7%) |
| Other | 301 | 3 (1.0%) | 3 (1.0%) | 4 (1.3%) | 291 (96.7%) |

**Table 4:** Number of respondents that believe they are suffering from a psychiatric diagnosis that has NOT been formally diagnosed by a healthcare professional

| **Diagnosis** | **N** | **Yes, currently** | **Yes, in the past** | **No** |
| --- | --- | --- | --- | --- |
| Depression | 395 | 78 (19.7%) | 33 (8.4%) | 284 (71.9%) |
| Anxiety | 390 | 120 (30.8%) | 26 (6.7%) | 244 (62.6%) |
| Panic disorder | 388 | 52 (13.4%) | 29 (7.5%) | 307 (79.1%) |
| Post-traumatic stress disorder | 391 | 78 (19.9%) | 27 (6.9%) | 286 (73.1%) |
| Obsessive compulsive disorder | 384 | 33 (8.6%) | 19 (4.9%) | 332 (86.5%) |
| Psychotic disorders | 382 | 8 (2.1%) | 4 (1.0%) | 370 (96.9%) |
| Bipolar disorder | 379 | 7 (1.8%) | 2 (0.5%) | 370 (97.6%) |
| Personality disorder | 383 | 22 (5.7%) | 15 (3.9%) | 346 (90.3%) |
| Impulse control disorder | 385 | 22 (5.7%) | 12 (3.1%) | 351 (91.2%) |
| Alcohol dependence | 386 | 9 (2.3%) | 11 (2.8%) | 366 (94.8%) |
| Drug dependence | 383 | 7 (1.8%) | 5 (1.3%) | 371 (96.9%) |
| Other | 309 | 10 (3.2%) | 3 (1.0%) | 296 (95.8%) |

When asked to rate the degree to which they felt their encephalitis was the cause of their mental health problems (0 = not at all, 100 = entirely), median response was 80.0 (IQR 50.0-100.0).

In total, 98/445 (22.0%) of respondents were currently under a community mental health team (secondary care), and 215/445 (48.3%) had undergone some form of talking therapy to address their mental health problems.

Of those who responded (n = 413), 24.2% felt they had not had access to appropriate healthcare for their mental health, and a further 29.3% thought the access to mental healthcare could have been better; 28.8% felt they had access to appropriate mental healthcare, and the remaining 17.7% had not looked (for breakdown by country please see Supplementary Information).

In total, 37.5% of respondents had thought about suicide, and 4.4% of all respondents had attempted suicide since their encephalitis diagnosis.

#### Psychiatric Diagnostic Screening Questionnaire

Using the cut-off scores provided in the PDSQ manual (see Supplementary information), participants met screening criteria in the following frequencies: social phobia 161/347 (46.4%), depression 143/347 (41.2%), OCD 136/346 (39.3%), PTSD 118/347 (34.0%), generalised anxiety disorder 111/347 (32.0%), agoraphobia 103/347 (29.7%), psychosis 88/347 (25.4%), panic disorder 84/347 (24.2%), alcohol use disorder 57/347 (16.4%), eating disorder 36/346 (10.4%), and drug use disorder 12/347 (3.5%).

The concordance of PDSQ scores versus self-reported diagnoses (formal diagnoses + self-reported missed diagnoses): depression 41.2% vs 44.4%; OCD 39.3% vs 14.9%; PTSD 34.0% vs 30.8%; GAD 24.2% vs 62.4%; psychosis 25.4% vs 3.9%; panic 24.2% vs 21.5%; alcohol use disorder 16.4% vs 4.1%; substance use disorder 3.5% vs 3.6%.

### Infectious versus autoimmune

Overall, rates of major self-reported psychiatric diagnoses and symptoms did not significantly differ between autoimmune and infectious encephalitis on Chi-square testing (*Table 5*).

**Table 5:**
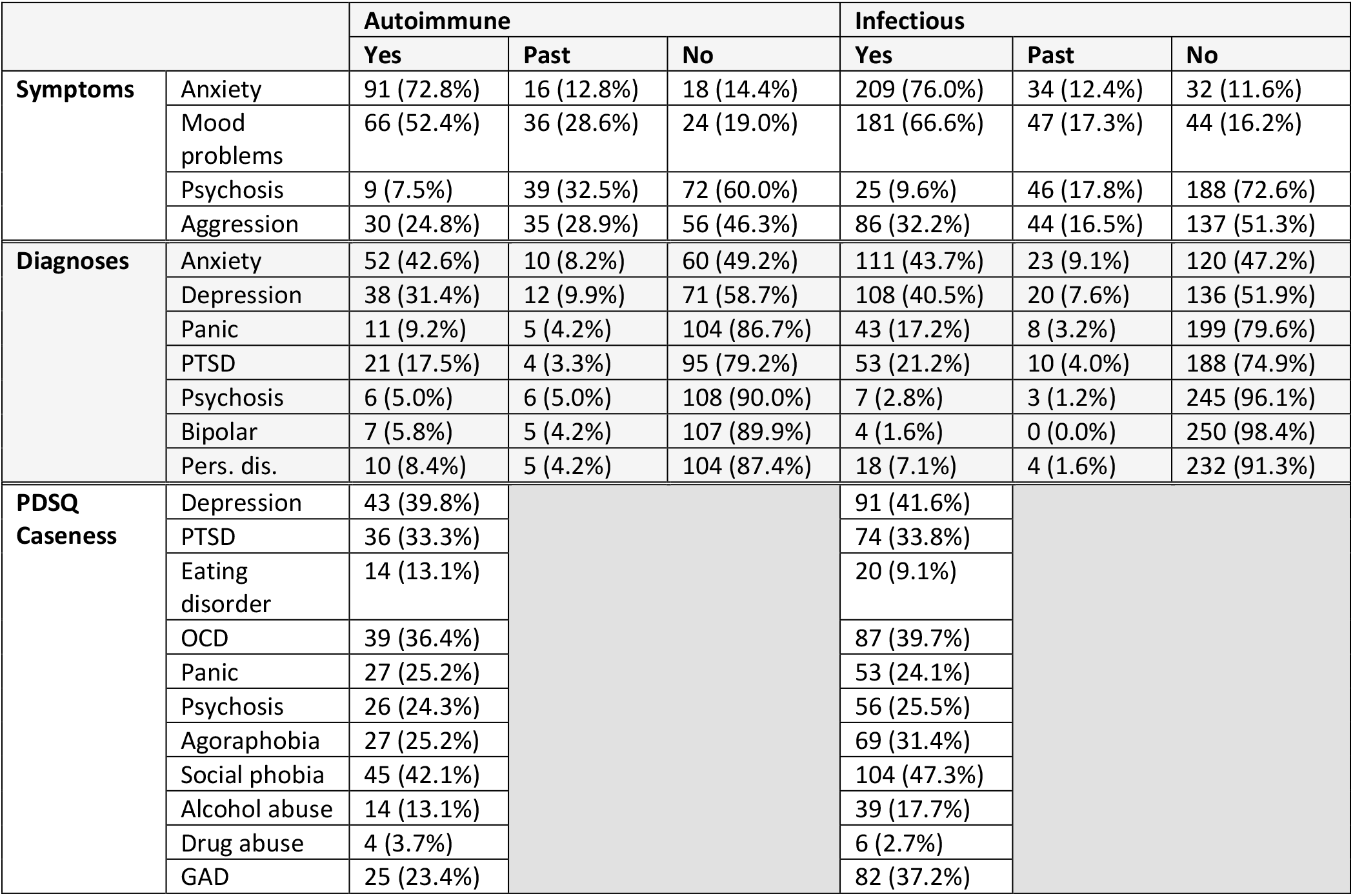
Comparison of psychiatric symptoms and diagnoses split by autoimmune and infectious subtypes.Pers. dis. = personality disorder.

### Brief Illness Perception Questionnaire

Mean and median scores on the BIPQ are summarised in *Table 6*.

**Table 6:**
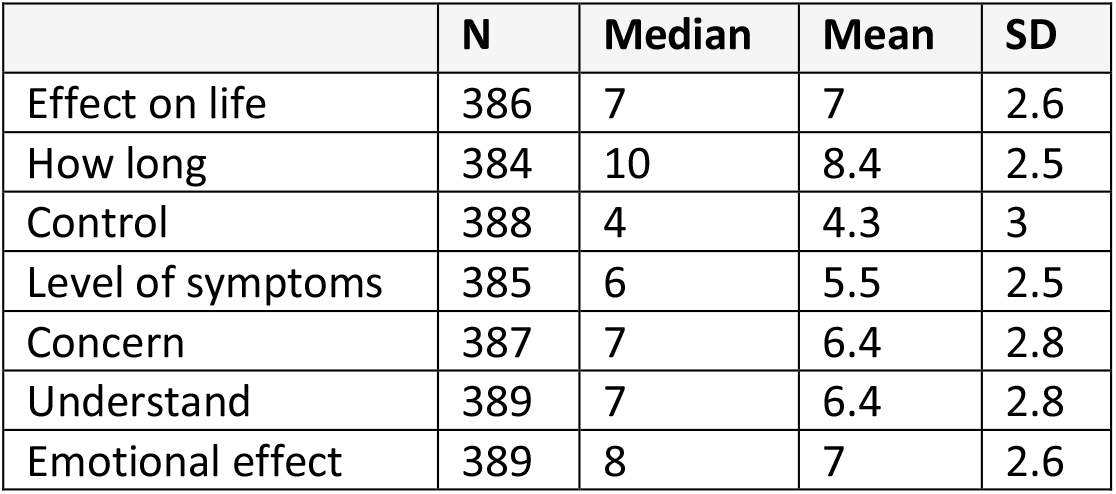
Median and mean from BIPQ. There were no differences in mean BIPQ subscores between autoimmune and infectious groups.

#### Maudsley Visual Analogue Scale

*Figure 3* summarises the results from the M3VAS. Mean (SD) values (/ 100) were as follows: suicidality 15.3 (26.6), mood 41.8 (32.1), anhedonia 47.3 (32.5); median scores: suicidality 0.0 (0.0 – 20.0), mood 40.0 (10.0 – 70.0), anhedonia 50.0 (19.0 – 77.5).

**Figure 3:**
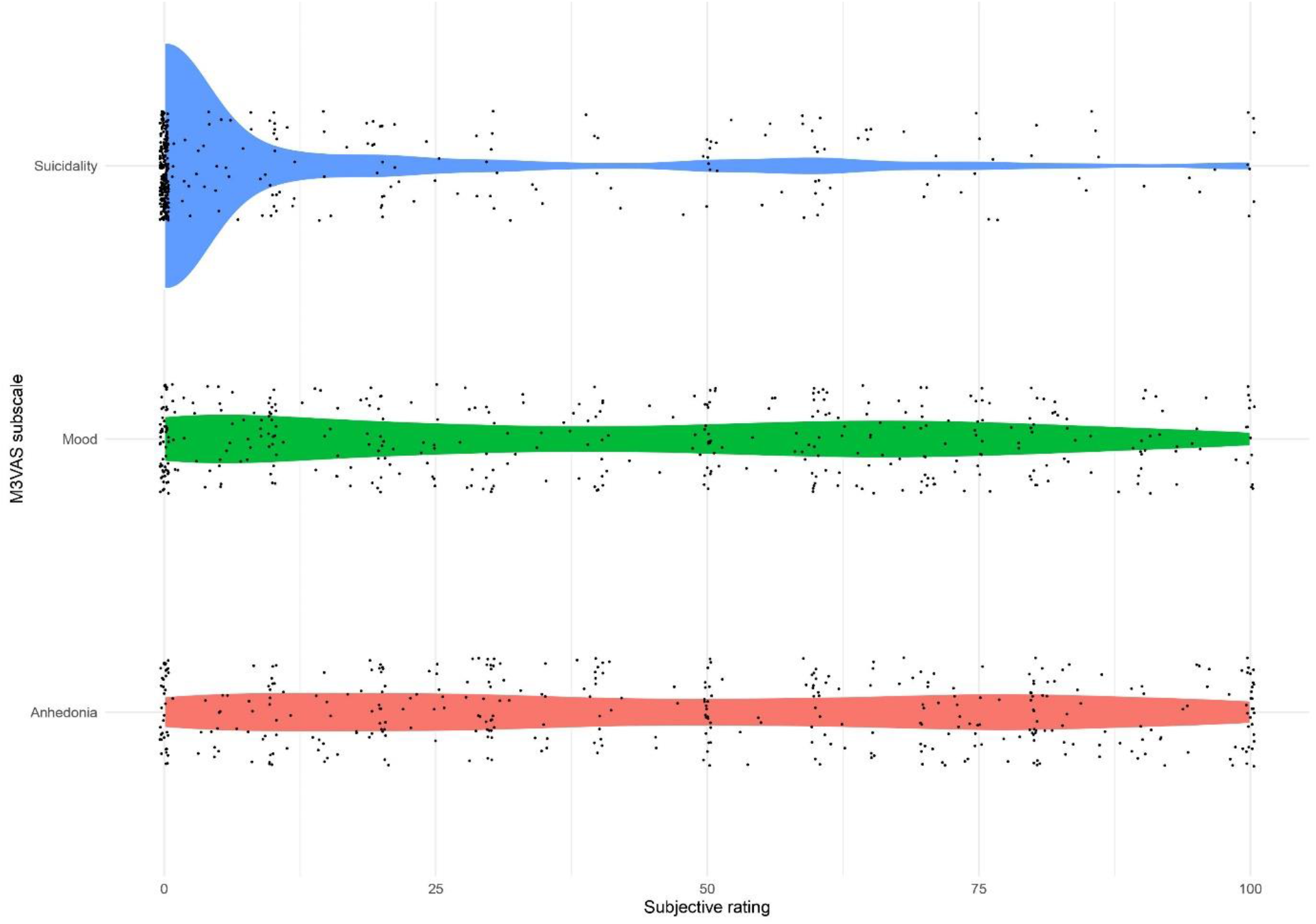
Violin plot of kernel density estimates from the M3VAS. Individual participant scores are represented by dots (the random vertical distribution of the dots is a visual aid only).

### Hypersensitivities

In total, 325/414 (78.5%) had experienced at least one hypersensitivity to sensory input; 99 (23.9%) had two hypersensitivities, and 164 (39.6%) experienced three or more hypersensitivities. Light hypersensitivities were reported by 233 (56.3%), sound hypersensitivities by 250 (60.4%), touch hypersensitivities by 93 (22.4%), taste hypersensitivities by 95 (22.9%), temperature hypersensitivities by 170 (41.1%), and smell hypersensitivities by 23 (5.6%).

Hypersensitivities were found to have a significant effect on daily life of respondents. On a scale of 0 = not affecting daily life at all, 10 = severely affecting daily life, light hypersensitivities were rated as mean 5.3 (SD 2.6), sound as mean 5.8 (SD 2.6), touch 5.5 (SD 2.7), taste 5.1 (2.8), and temperature 5.8 (2.5). There were no differences between anxiety & depression scores in those with or without hypersensitivities.

### Management of symptoms

Overall, respondents reported that they had received a variety of treatments for the management of their symptoms following encephalitis. The most common treatment amongst respondents was the use of medication (77.2%, n = 287); more specifically, non-opiate painkillers (60.1%, n = 140) antidepressants (59.7%, n = 148), benzodiazepines (41.3%, n = 92), sleeping tablets (34.5%, n = 75), opiates (33.0%, n = 72), antipsychotics (23.2%, n = 49), and medicinal cannabis (13.7%) had been used. Self-reported effectiveness of these treatments are summarised in *Table 7*.

**Table 7:** Self-reported effectiveness of treatments.

| Medication | Unhelpful | Somewhat helpful | Helpful |
| --- | --- | --- | --- |
| Antidepressants ($n = 126$) | 33 (26.2%) | 47 (37.3%) | 46 (36.5%) |
| Benzodiazepines ($n = 81$) | 15 (18.5%) | 28 (34.6%) | 38 (46.9%) |
| Antipsychotics ($n = 44$) | 19 (43.2%) | 13 (29.5%) | 12 (27.3%) |
| Sleeping tablets ($n = 71$) | 16 (22.5%) | 33 (46.5%) | 22 (31.0%) |
| Opiates ($n = 64$) | 12 (18.8%) | 28 (43.8%) | 24 (37.5%) |
| Non-opioid painkillers ($n = 114$) | 25 (21.9%) | 40 (35.1%) | 49 (43.0%) |
| Medicinal cannabis ($n = 77$) | 13 (16.9%) | 14 (18.4%) | 50 (64.9%) |

Other forms of therapy received in respondents include occupational therapy (n= 143, 40.4%), neuropsychological rehabilitation (n = 132, 37.3%), cognitive behavioural therapy (n = 123, 34.1%), and other psychotherapy (n = 107, 30.6%).

Legal non-prescribed substances had also been used to help manage post-encephalitis symptoms in 19.1% of respondents and include energy drinks/caffeine (n = 27, 7.4%), CBD (n = 27, 7.4%), alcohol (n = 19, 5.2%), tobacco (n = 10, 2.7%), and e-cigarettes/nicotine (n = 4, 1.1%). Mean (SD) effectiveness of legal substances were as follows (0 = not effective at all, 100 = completely effective): caffeine 53.6 (32.0), CBD 67.3 (29.9), alcohol 42.3 (23.8), tobacco 49.7 (40.9), an e-cigarettes/nicotine 49.7 (47.6).

In total, 6.5% of respondents had obtained medication without prescription to treat their encephalitis.

In total, 7.5% (n = 29) of respondents had tried illegal street substances to help manage post-encephalitis symptoms and included cannabis (n = 21, 5.4%), psilocybin (n = 4, 1.0%), and cocaine (n = 3, 0.8%).

## Discussion

This large international survey on psychiatric symptoms following encephalitis represents the largest such study to date, acquiring data on an unprecedented breadth of psychiatric symptomatology in a markedly under-researched patient group. Responses from participants indicated high rates of self-reported mental health symptoms and diagnoses, with many reporting that they had not received adequate psychiatric care following their encephalitis.

The psychiatric community has become increasingly aware of encephalitis since the first descriptions of NMDAR antibody encephalitis presenting with florid psychotic symptoms, and increasingly autoimmune encephalitis is on the list of differential diagnoses for acute, new-onset severe psychiatric presentations including psychosis [22, 23]. The current study found that psychiatric symptoms are also very common in the so-called ‘recovery’ phases of encephalitis, indicating that awareness of this disorder among mental health professionals should not be restricted to acute presentations only.

The relationship between mental and physical health in neurological disorders is complex and bidirectional: neurology patients with emotional disorders including depression and anxiety report a higher burden of somatic symptoms [24] and recovery from depression in neurology patients is associated with improvement in overall health status, including on physical health-related subscales [25]. There are many data that indicate that neurological disorders are seen with comorbid psychiatric disorders at rates much higher than population averages, with just a few examples including epilepsy, Parkinson’s disease, and migraine [26]. Traumatic brain injury, another form of acquired brain injury, is associated with the development of both neurological and psychiatric sequelae [27].

While the issue has not been systematically assessed in many studies, the degree of psychiatric morbidity indicated by the current survey provides a strong *prima facie* argument for the continued involvement of mental health services in the long-term follow-up of patients who have had encephalitis. Numerous studies have shown that physicians in general and neurologists in particular do not systematically ask about many aspects of patient’s mental health in routine outpatient clinical encounters and as such considerable mental health difficulties may be missed [24]. In a 2022 survey of 5593 adults living in England with neurological disorders, 61% reported not being asked about their mental wellbeing by a health or social care professional within the last three years [28].

Therefore, in line with recent recommendations that liaison psychiatry should adopt proactive models of working rather than the reactive approach taken in many centres currently [29], we suggest that mental health outcomes could be improved in encephalitis either by the use of routine screening instruments in neurology-led clinical encounters, or the ongoing involvement of psychiatry or neuropsychiatry in the clinical follow-up of these patients. In one study of long-term psychosocial outcomes in 61 patients with anti-NMDAR encephalitis, involvement of psychiatry was associated with an eight-fold increased odds of a patient returning to work or education [16]. A broader systematic evaluation of the efficacy of mental health interventions in patients with encephalitis-associated mental illness is urgently required.

Our data regarding suicidality and suicide following encephalitis are deeply concerning. While suicidality and indeed completed suicide has been observed as a feature of anti-NMDA receptor encephalitis in one Chinese series [30], this represents a relatively understudied area given the potential contribution of suicide to avoidable deaths in this disorder. In a pertinent epidemiological study of over 7 million Danish citizens, patients who had had encephalitis were at an increased risk of death by suicide (incident rate ratio 1.7 compared to people with no neurological diagnosis; 95% CI, 1.3-2.3), a rate comparable to individuals with epilepsy, Parkinson’s disease and head injury, and higher than stroke and dementia [31]. This is a clinical concern of considerable urgency and mandates further research as well as increased clinical awareness amongst all teams providing health and social care to patients who have experienced encephalitis.

The high rates of sensory hypersensitivities in people who have had encephalitis has not previously been reported. Sensory processing issues are common in autism spectrum disorders, although the underlying neurobiology has not been elucidated [32]. Visual and auditory hypersensitivities have been reported in patients with traumatic brain injury (mainly mild), where their presence correlates with mental distress [33] and a poorer quality of life [34], and in brain tumours where their presence may be associated with depressive symptoms [35]. Furthermore, sensory hypersensitivities, particularly towards light, sound, and touch, are seen very commonly in migraine with and without aura [36]. Further work is required to more fully elucidate the nature, extent, underlying mechanisms, and impacts of sensory hypersensitivities in people who have had encephalitis.

Misdiagnosis was commonly reported by respondents. In this study, a physical health misdiagnosis was more common than a psychiatric misdiagnosis, likely because of the predominance of infectious encephalitis diagnoses among respondents. Increasing the awareness and understanding of encephalitis among healthcare professionals is therefore likely to be useful in the general hospital/general practice as well as the mental health setting. Misdiagnosis is bidirectional; a recent study indicated that misdiagnosis occurs in around one quarter of patients with autoimmune encephalitis, albeit with significant variance between healthcare settings. The most common conditions incorrectly diagnosed as autoimmune encephalitis were functional neurologic disorder (FND), neurodegenerative disease, and primary psychiatric disease [7]. FND in particular is likely to be a common misdiagnosis in both directions and may be seen as an additional diagnosis in some cases.

The high rates of psychiatric disorders following encephalitis very likely have multiple overlapping causal factors, which include, but are not limited to, structural or functional brain alterations which are a direct result of the pathogenic infiltration or immune response, medication response (e.g., to steroids), residual symptoms following delirium or ITU stay (for example) in those with a severe acute illness, psychological readjustment following the experience of a life-threatening illness, the repercussions of change in functional or social status following a significant illness, and the effects of resultant physical disability. This survey was not equipped to answer aetiological questions on the nature of psychiatric disorder aetiology following encephalitis; this will be a target for future research, as insights might help shape the evidence-based development of preventative and therapeutic approaches to encephalitis-associated mental illness (including ongoing work from our group in developing cognitive-behavioural models to aid with the psychological support of this patient group).

### Strengths and limitations

Strengths of this study include the large international dataset, broad focus, and the emphasis on patients’ own perspectives. The use of an online platform to complete the questionnaire meant that it was possible to access patients across the world, who may not have been able to travel for research purposes, for example, due to distance, travel restrictions, symptoms, financial barriers.

Nevertheless, the study has a number of limitations. The responses to the questionnaire were not clinically validated, and we relied on self-report. The data is likely to be subject to both response bias, whereby those with active psychiatric symptoms are more likely to undertake the survey in the first place, and recall bias, where respondents are biased towards recalling symptoms due to the prompting inherent in the questioning. Nevertheless, conversely, those with more severe symptoms may be prevented from completing the survey, thus underestimating symptom burden.

There are attendant limitations to using screening tools such as the PDSQ, which may over-estimate the prevalence of included disorders; these tools are also limited in the range of disorders that they are equipped to capture, therefore other disorders may be missed. The positive predictive value of the PDSQ is, for some disorders, low.

This study recruited voluntary participants through a global patient organisation, the Encephalitis Society, specifically through their social media platforms and mailing lists. This may not be representative of the entire encephalitis population, insofar as more severely physically or cognitively challenged people may have been less able to complete the survey, despite our efforts to counterbalance digital exclusion by facilitating carers to complete it on their behalf. Conversely, however, some individuals who have made a full recovery may be less likely to engage with the Encephalitis Society or with encephalitis-related content online, and therefore potentially less likely to access or complete our questionnaire. Finally, the questionnaire was only available in English, and hence is biased towards English-speaking populations.

## Conclusions

Overall, the large international survey indicates that psychiatric symptoms following encephalitis are common. Despite the high rates, many respondents reported either that they had not been diagnosed with a mental health problem they felt that they had, and/or that mental healthcare provision following their encephalitis was inadequate. Limitations to this survey include respondent and recall bias, and generalisability is limited by the online sample.

Overall, these results highlight a need for increased provision of proactive psychiatric care for these patients and represent a call to action for increased research on mental health outcomes of encephalitis so that this patient group can be better supported. Given the treatment-responsiveness of many mental health symptoms and diagnoses, this is likely to represent a global opportunity for reducing morbidity and mortality in this challenging condition.

## Data Availability

Data available upon reasonable request (post-full publication).

## References

1. Encephalitis Society (2022) Encephalitis: an in-depth review and gap analysis of key variables affecting global disease burden..

2. Tunkel AR, Glaser CA, Bloch KC, Sejvar JJ, Marra CM, Roos KL, Hartman BJ, Kaplan SL, Scheld WM, Whitley RJ (2008) The management of encephalitis: clinical practice guidelines by the Infectious Diseases Society of America. Clin Infect Dis 303–327.

3. Encephalitis Society (2018) Encephalitis in adults: a guide. 9.

4. Granerod J, Ambrose HE, Davies NWS, Clewley JP, Walsh AL, Morgan D, Cunningham R, Zuckerman M, Mutton KJ, Solomon T (2010) Causes of encephalitis and differences in their clinical presentations in England: a multicentre, population-based prospective study. Lancet Infect Dis 10:835–844.

5. Titulaer MJ, McCracken L, Gabilondo I, et al (2013) Treatment and prognostic factors for long-term outcome in patients with anti-NMDA receptor encephalitis: an observational cohort study. Lancet Neurol 12:157–165.

6. Gibson LL, Pollak TA, Blackman G, Thornton M, Moran N, David AS (2018) The Psychiatric Phenotype of Anti-NMDA Receptor Encephalitis. J Neuropsychiatry Clin Neurosci 31:70–79.

7. Flanagan EP, Geschwind MD, Lopez-Chiriboga AS, et al (2023) Autoimmune Encephalitis Misdiagnosis in Adults. JAMA Neurol 80:30–39.

8. Clarke M, Newton RW, Klapper PE, Sutcliffe H, Laing I, Wallace G (2006) Childhood encephalopathy: viruses, immune response, and outcome. Dev Med Child Neurol 48:294– 300.

9. Arciniegas DB, Anderson CA (2004) Viral encephalitis: neuropsychiatric and neurobehavioral aspects. Curr Psychiatry Rep 6:372–379.

10. Whitley RJ (1990) Viral encephalitis. N Engl J Med 323:242–250.

11. Finke C, Kopp UA, Prüss H, Dalmau J, Wandinger K-P, Ploner CJ (2012) Cognitive deficits following anti-NMDA receptor encephalitis. J Neurol Neurosurg Psychiatry 83:195–198.

12. McKeon GL, Robinson GA, Ryan AE, Blum S, Gillis D, Finke C, Scott JG (2018) Cognitive outcomes following anti-N-methyl-D-aspartate receptor encephalitis: a systematic review. J Clin Exp Neuropsychol 40:234–252.

13. Liu X, Zhang L, Chen C, Gong X, Lin J, An D, Zhou D, Hong Z (2019) Long-term cognitive and neuropsychiatric outcomes in patients with anti-NMDAR encephalitis. Acta Neurol Scand 140:414–421.

14. Chou IC, Lin CC, Kao CH (2015) Enterovirus encephalitis increases the risk of attention deficit hyperactivity disorder: A Taiwanese population-based case-control study. Med (United States) 94:1–5.

15. Harris L, Griem J, Gummery A, et al (2020) Neuropsychological and psychiatric outcomes in encephalitis: A multi-centre case-control study. PLoS One 15:1–24.

16. Blum RA, Tomlinson AR, Jetté N, Kwon CS, Easton A, Yeshokumar AK (2020) Assessment of long-term psychosocial outcomes in anti-NMDA receptor encephalitis. Epilepsy Behav 108:10–12.

17. Mailles A, De Broucker T, Costanzo P, Martinez-Almoyna L, Vaillant V, Stahl JP (2012) Long-term outcome of patients presenting with acute infectious encephalitis of various causes in France. Clin Infect Dis 54:1455–1464.

18. Ng BY, Lim CCT, Yeoh A, Lee WL (2004) Neuropsychiatrie sequelae of Nipah virus encephalitis. J Neuropsychiatry Clin Neurosci 16:500–504.

19. Moulton CD, Strawbridge R, Tsapekos D, Oprea E, Carter B, Hayes C, Cleare AJ, Marwood L, Mantingh T, Young AH (2021) The Maudsley 3-item Visual Analogue Scale (M3VAS): Validation of a scale measuring core symptoms of depression. J Affect Disord 282:280–283.

20. Broadbent E, Petrie KJ, Main J, Weinman J (2006) The Brief Illness Perception Questionnaire. J Psychosom Res 60:631–637.

21. Zimmerman M, Mattia JI (1999) The reliability and validity of a screening questionnaire for 13 DSM-IV Axis I disorders (the Psychiatric Diagnostic Screening Questionnaire) in psychiatric outpatients. J Clin Psychiatry 60:677–683.

22. Ariño H, Coutinho E, Pollak TA, Stewart R (2021) Real-world experience of assessing antibodies against the N-methyl-D-aspartate receptor (NMDAR-IgG) in psychiatric patients. A retrospective single-centre study. Brain Behav Immun 98:330–336.

23. Pollak TA, Lennox BR, Müller S, et al (2020) Autoimmune psychosis: an international consensus on an approach to the diagnosis and management of psychosis of suspected autoimmune origin. The Lancet Psychiatry 7:93–108.

24. Carson AJ, Ringbauer B, MacKenzie L, Warlow C, Sharpe M (2000) Neurological disease, emotional disorder, and disability: they are related: a study of 300 consecutive new referrals to a neurology outpatient department. J Neurol Neurosurg & Psychiatry 68:202 LP – 206.

25. Carson AJ, Postma K, Stone J, Warlow C, Sharpe M (2003) The outcome of depressive disorders in neurology patients: a prospective cohort study. J Neurol Neurosurg & Psychiatry 74:893 LP – 896.

26. Hesdorffer DC (2016) Comorbidity between neurological illness and psychiatric disorders. CNS Spectr 21:230–238.

27. Perry DC, Sturm VE, Peterson MJ, et al (2016) Association of traumatic brain injury with subsequent neurological and psychiatric disease: a meta-analysis. J Neurosurg JNS 124:511–526.

28. Neurological Alliance (2022) Together for the 1 in 6: England findings from My Neuro Survey.

29. Sharpe M, Toynbee M, Walker J (2020) Proactive Integrated Consultation-Liaison Psychiatry: A new service model for the psychiatric care of general hospital inpatients. Gen Hosp Psychiatry 66:9–15.

30. Zhang L, Sander JW, Zhang L, et al (2017) Suicidality is a common and serious feature of anti-N-methyl-D-aspartate receptor encephalitis. J Neurol 264:2378–2386.

31. Erlangsen A, Stenager E, Conwell Y, Andersen PK, Hawton K, Benros ME, Nordentoft M, Stenager E (2020) Association Between Neurological Disorders and Death by Suicide in Denmark. JAMA 323:444–454.

32. Robertson CE, Baron-Cohen S (2017) Sensory perception in autism. Nat Rev Neurosci 18:671– 684.

33. Shepherd D, Landon J, Kalloor M, Theadom A (2019) Clinical correlates of noise sensitivity in patients with acute TBI. Brain Inj 33:1050–1058.

34. Shepherd D, Landon J, Kalloor M, Barker-Collo S, Starkey N, Jones K, Ameratunga S, Theadom A, Group BR (2020) The association between health-related quality of life and noise or light sensitivity in survivors of a mild traumatic brain injury. Qual Life Res 29:665–672.

35. Ochi R, Saito S, Hiromitsu K, Shigemune Y, Shinoura N, Yamada R, Midorikawa A (2022) Sensory hypo- and hypersensitivity in patients with brain tumors. Brain Inj 36:1053–1058.

36. Pearl TA, Dumkrieger G, Chong CD, Dodick DW, Schwedt TJ (2020) Sensory Hypersensitivity Symptoms in Migraine With vs Without Aura: Results From the American Registry for Migraine Research. Headache J Head Face Pain 60:506–514.

